# Hospitalization and 30-day fatality in 121,263 COVID-19 outpatient cases

**DOI:** 10.1101/2020.05.04.20090050

**Authors:** Daniel Prieto-Alhambra, Elisabet Balló, Ermengol Coma, Núria Mora, María Aragón, Albert Prats-Uribe, Francesc Fina, Mència Benítez, Carolina Guiriguet, Mireia Fàbregas, Manuel Medina-Peralta, Talita Duarte-Salles

**Affiliations:** Fundació Institut Universitari per a la recerca a l’Atenció Primària de Salut Jordi Gol i Gurina (IDIAPJGol), Barcelona, Spain; Centre for Statistics in Medicine, NDORMS, University of Oxford; Sistemes d’Informació dels Serveis d’Atenció Primària (SISAP), Institut Català de la Salut (ICS), Barcelona, Spain; Equip d’Atenció Primària de Salt, Institut Català de la Salut, Girona, Spain; Equip d’Atenció Primària Gòtic, Institut Català de la Salut, Barcelona, Spain

**Keywords:** COVID-19, Epidemiology, coronavirus, fatality, hospital admission

## Abstract

**Background:** To date, characterisation studies of COVID-19 have focussed on hospitalised or intensive care patients. We report for the first time on the natural history of COVID-19 disease from first diagnosis, including both outpatient and hospital care.

**Methods:** Data was obtained from SIDIAP, a primary care records database covering >6 million people (>80% of the population of Catalonia), linked to COVID-19 RT-PCR tests, hospital emergency and inpatient, and mortality registers. All participants >=15 years, diagnosed with COVID-19 in outpatient between 15th March and 24th April 2020 (10^th^ April for outcome studies) were included. Baseline characteristics, testing, and 30-day outcomes (hospitalisation for COVID-19 and all-cause fatality) were analysed.

**Results:** A total of 121,263 and 95,467 COVID-19 patients were identified for characterisation and outcome studies, respectively. Women (57.8%) and age 45-54 (20.2%) were predominant. 44,709 were tested, with 32,976 (73.8%) PCR+. From 95,467 cases, a 14.6% [14.4–14.9] were hospitalised in the month after diagnosis, with male predominance (19.2% vs 11.3%), peaking at age 75-84. Overall 30-day fatality was 4.0% [95%CI 3.9%-4.2%], higher in men (4.8%) than women (3.4%), increasing with age, and highest in those residing in nursing homes (25.3% [24.2% to 26.4%]).

**Conclusions:** COVID-19 is seen in all age-sex strata, but severe forms of disease cluster in older men and nursing home residents. Although initially managed in primary care, 15% of cases require hospitalization within a month, with overall fatality of 4%.

## Introduction

COVID-19 started as an outbreak in Wuhan, China, in December 2019, and rapidly developed into a global pandemic, causing substantial morbidity and fatality burden and straining healthcare systems worldwide^1^. In Europe, it was first reported in late January, with the first case in Spain reported a month later, although some studies suggest community transmission already during that time^2^. To date, Spain has reported the second-highest number of confirmed cases and the fourth death toll of COVID-19 in the world.

Most COVID-19 patients present influenza-like symptoms, including fever, dry cough, fatigue, or sore throat^3^, and are therefore eligible for outpatient or primary care management in first instance. Despite this, studies to date have focussed on the characteristics and prognosis of COVID-19 patients in hospitalised^4^ or intensive care patients^5^, skewing current estimates of the morbi-mortality of COVID-19 globally. It is therefore essential to characterise patients from their first diagnosis in order to achieve a more complete understanding of the prognosis of this disease. Healthcare systems of universal coverage like the Spanish or the UK rely on general practitioners (GPs) as gatekeepers, with all patients seen in primary care before admission to hospital^6^. Primary care electronic health records from such healthcare systems, when linked to hospital admissions and mortality registers, offer therefore a unique opportunity to fully characterise the natural history of COVID-19.

We obtained primary care records for a large number of COVID-19 cases, linked to reverse transcriptase–polymerase chain reaction (RT-PCR) data, and hospital and mortality registers to describe the natural history of COVID-19 in Catalonia, Spain. First, we characterised the socio-demographics, comorbidities, and medicines used at the time of diagnosis. Secondly, we studied the prognosis of COVID-19 by ascertaining 30-day hospital admissions and all-cause fatality from outpatient and primary care diagnosis.

## Methods

### Study design and data sources

We performed a cohort study with prospectively collected data from the Information System for Research in Primary Care (SIDIAP; www.sidiap.org) in Catalonia, Spain^7^. SIDIAP contains anonymized primary care electronic health records for over six million people, covering a representative >80% of the Catalan population, since 2006. SIDIAP includes high-quality and validated diagnoses (International Classification for Diseases, 10th revision, Clinical Modification [ICD-10-CM]), medicine prescriptions, laboratory tests, and lifestyle and socio-demographics^8,9^. For this particular study, SIDIAP was further linked to the region-wide population-based hospital and outpatient emergency register^10^, to the bespoke central database of RT-PCR COVID-19 tests, and to the regional mortality registry.

### Study participants

We included all individuals aged 15 or older with COVID-19 identified by a positive RT-PCR test for Severe acute respiratory syndrome coronavirus 2 (SARS-CoV-2) and/or a clinical diagnosis (ICD-10-CM codes B34.2, B97.21, B97.29, J12.81) recorded in primary care from March 15th, 2020 to April 24th, 2020. We excluded individuals who were hospitalised with COVID-19 before their index date (prevalent cases). For the analysis of outcomes (hospital admission or fatality), only subjects with an index date before April 10th 2020 were included to guarantee at least 14 days of follow-up from index date. In addition, subjects with a prevalent diagnosis of pneumonia and/or a hospital admission for respiratory symptoms subsequently diagnosed with COVID-19 were excluded from the analyses of hospital admissions (but not from the analysis of fatality) to avoid misclassification.

Participants were followed from the earliest of a first RT-PCR+ or clinical diagnosis (index date) until death or end of study period (April 24th, 2020). Repeat RT-PCR testing was dismissed.

### Baseline characteristics and comorbidities

Socio-demographics were assessed at index date, to include sex, age (in years), place of residence (community vs nursing home), rurality (rural, urban), and nationality (Spain, other). Rural areas were defined as areas with less than 10,000 inhabitants and a population density lower than 150 inhabitants/km^2^. We assessed socio-economic status using the validated MEDEA deprivation index, previously linked to SIDIAP^11^, calculated by census tract level in urban areas and categorised in quartiles where 1st and 4th quartiles are least and most deprived areas, respectively. Rural areas were categorized separately.

We defined pre-existing comorbidities as the presence of a diagnosis code recorded any time prior to index date and still active at the time of COVID-19 diagnosis for a pre-specified list of conditions based on previous literature^3–5^. Lists of ICD-10-CM codes for each of these conditions are provided in *Supplementary Table 1*.

Finally, we characterised use of long-term medicines based on primary care prescriptions active at the time of diagnosis (index date). A pre-specified list of medicines was created based on the same previous literature^3–5^, and identified using ATC codes (*Supplementary Table 2*).

### Outcomes

Both study outcomes were studied in the 30 days after index date. Two outcomes were studied, namely: COVID-19 related hospital admission, and all-cause death. Hospitalisations were ascertained from linked hospital data covering emergency rooms and inpatient administrative data for the whole region of Catalonia. COVID-19 related admissions were identified using a bespoke list of diagnostic codes as recorded from hospital discharge data (ICD-10-CM codes listed in *Supplementary Table 3*). Date of death was obtained from linked regional mortality data.

### Statistical analysis

For descriptive analyses, median (inter-quartile range) or n (%) were reported for each of the patient characteristics detailed above. Cumulative n and % for hospital admissions and fatality were reported as obtained from the data, whilst 30-day probabilities were obtained from Kaplan-Meier estimates, stratified by sex, age (10-year bands), nursing home residence status, and RT-PCR result where available. Kaplan-Meier estimates were depicted for both study outcomes stratified by sex and age group. All analyses were conducted using R version 3.5.1.

## Results

Overall, 121,263 cases of COVID-19 were identified and eligible for descriptive analyses. Of these, 46,674 (38.5%) had a RT-PCR test result within a median (inter-quartile range) of 3.7 (7.4) days: 34,254 (73.4%) were positive, 11,108 (23.8%) negative, and the remaining 1,312 (2.8%) were inconclusive.

A total of 98,004/121,263 (80.8%) had 14 days or more of follow-up available and were therefore included for fatality analyses, while 95,467/121,263 (78.7%) were included for the analysis of hospital admissions. A population flow-chart is provided in Figure 1.

**Figure 1.**
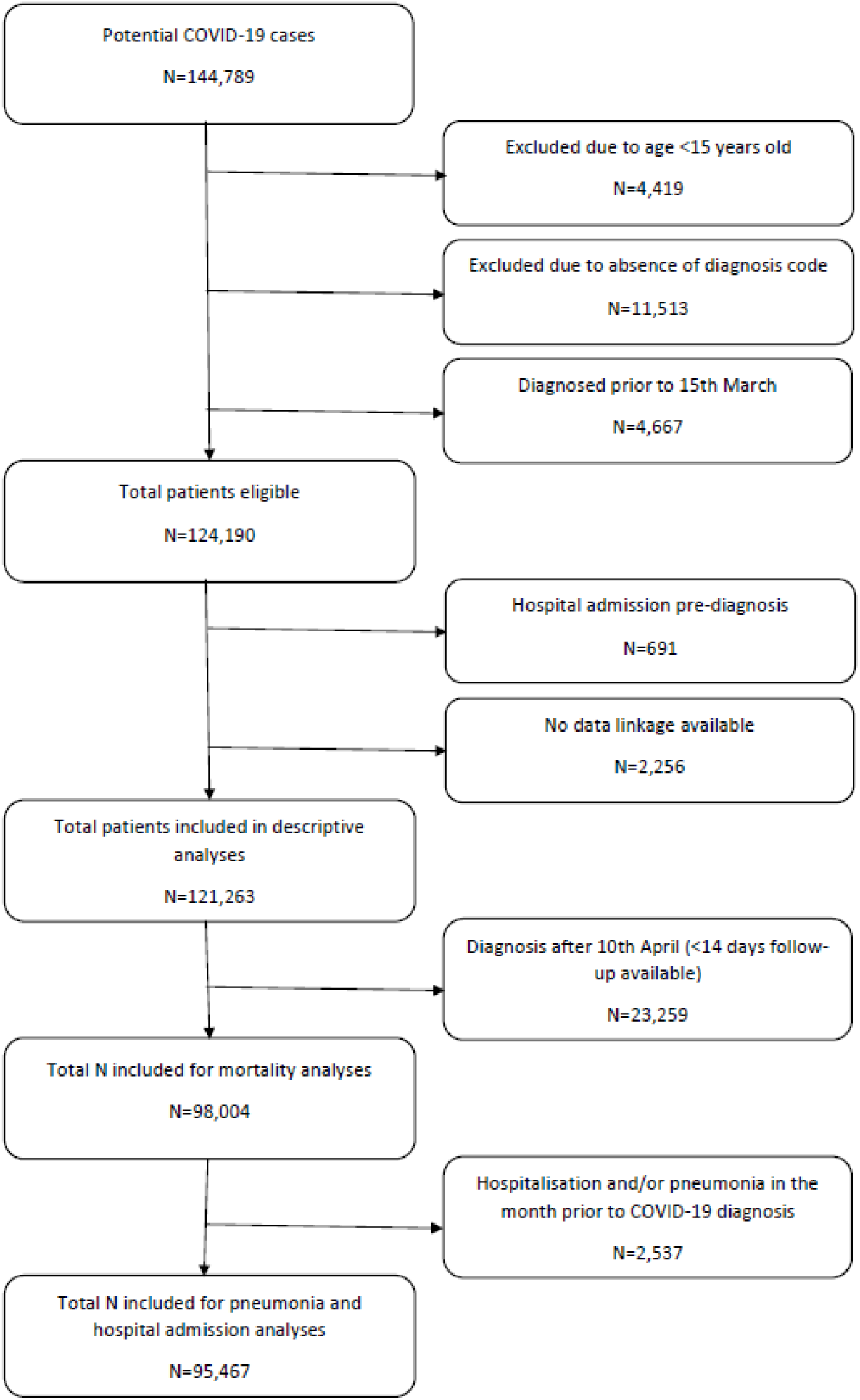
Population Flow-chart.

Table 1 summarizes the baseline characteristics of participants at the time of COVID-19 diagnosis. COVID-19 infectees were predominantly women (58.3%) in their middle ages (20.8% 35-44, 20.8% 45-54, and 15.4% 55-64 years old). A 21.0% (25,478) of infected people lived in urban most deprived areas, and an additional 17.5% (21,203) in rural areas. A substantial 10,795 cases (8.9% of total) were identified amongst subjects living in nursing homes, and a total of 592 (0.5% of total) women were pregnant at the time of COVID-19 diagnosis.

**Table 1.** Baseline characteristics at the time of COVID-19 diagnosis^*^.

| Variable | Value | N | % |
| --- | --- | --- | --- |
| ***Socio-demographics*** |  |  |  |
| Age | 15 to 24 years old | 6,890 | 5.68% |
| Age | 25 to 34 years old | 16,048 | 13.23% |
| Age | 35 to 44 years old | 25,197 | 20.78% |
| Age | 45 to 54 years old | 25,257 | 20.83% |
| Age | 55 to 64 years old | 18,681 | 15.41% |
| Age | 65 to 74 years old | 9,931 | 8.19% |
| Age | 75 to 84 years old | 9,034 | 7.45% |
| Age | 85 or older | 10,225 | 8.43% |
| Sex | Women | 70,731 | 58.33% |
| Nationality | Spain | 107,231 | 88.43% |
| Nationality | Other | 14,032 | 11.57% |
| Socio-economic status | Urban - Quartile 1 (Least deprived) | 30,718 | 25.33% |
| Socio-economic status | Urban - Quartile 2 | 18,017 | 14.86% |
| Socio-economic status | Urban - Quartile 3 | 21,564 | 17.78% |
| Socio-economic status | Urban - Quartile 4 (Most deprived) | 25,478 | 21.01% |
| Socio-economic status | Rural | 21,203 | 17.49% |
| Socio-economic status | Missing | 4,283 | 3.53% |
| Residence | Nursing home | 10,795 | 8.90% |
| Pregnancy | Yes | 592 | 0.49% |
| ***Comorbidities*** |  |  |  |
| Atrial Fibrillation |  | 4,661 | 3.84% |
| Osteoarthritis |  | 18,621 | 15.36% |
| Asthma |  | 8,260 | 6.81% |
| Ischaemic heart disease |  | 3,739 | 3.08% |
| Dementia |  | 5,515 | 4.55% |
| Diabetes mellitus |  | 11,829 | 9.75% |
| Liver disease |  | 5,565 | 4.59% |
| Hypertension |  | 29,417 | 24.26% |
| Heart Failure |  | 3,006 | 2.48% |
| Cerebrovascular disease |  | 2,835 | 2.34% |
| COPD |  | 3,914 | 3.23% |
| Chronic kidney disease |  | 6,554 | 5.40% |
| Cancer (all except non-melanoma skin cancer) |  | 9,032 | 7.44% |
| Obesity |  | 24,132 | 19.90% |
| HIV infection |  | 311 | 0.26% |
| Hepatitis B |  | 319 | 0.26% |
| Hepatitis C |  | 760 | 0.63% |
| ***Long-term medicine/s use*** |  |  |  |
| Analgesics |  | 26,651 | 21.98% |
| Sedatives/hypnotics |  | 20,529 | 16.93% |
| Anticoagulants |  | 16,423 | 13.54% |
| Antidepressants |  | 19,647 | 16.20% |
| Antiepileptics |  | 7,749 | 6.00% |
| Antipsychotics |  | 7,885 | 6.50% |
| PPI/antiacids |  | 23,079 | 19.03% |
| Systemic corticosteroids |  | 2,746 | 2.26% |
| Oral anti-diabetics |  | 8,504 | 7.01% |
| Insulin |  | 3,616 | 2.98% |
| Lipid modifying agents |  | 15,366 | 12.67% |
| COPD/asthma inhalers |  | 13,012 | 10.73% |
| Alpha-blockers |  | 717 | 0.59% |
| Beta-blockers |  | 8,646 | 7.13% |
| Calcium channel blockers |  | 6,966 | 5.74% |
| Diuretics |  | 10,158 | 8.38% |
| ACEi/ARBs |  | 14,225 | 11.73% |
| Combination antihypertensives |  | 6,560 | 5.41% |

\*Data are for persons with a RT-PCR test positive for Severe acute respiratory syndrome coronavirus 2 (SARS-CoV-2) and/or a clinical diagnosis recorded in primary care from March 15th, 2020 to April 24th, 2020.

Most common comorbidities included hypertension (24.3%), obesity (19.9%) and osteoarthritis (15.4%), whilst least common ones were chronic viral hepatitis (0.6% type C, 0.3% type B) and HIV (0.3%). The top 10 most commonly used long-term therapies at the time of COVID-19 diagnosis were analgesics (22.0%), PPI/antiacids (19.0%), sedatives/hypnotics (16.9%), antidepressants (16.2%), antithrombotics (13.5%), lipid modifying agents (12.7%), ACEi/ARBs (11.7%), chronic obstructive pulmonary disease (COPD)/asthma inhalers (10.7%), diuretics (8.4%), and beta blockers (7.1%).

A total of 14,141/95,467 patients were hospitalized for complications of COVID-19, equivalent to a cumulative incidence of 14.6% [14.4% to 14.9%] in the month after diagnosis, higher amongst men at 19.2% [18.9% to 19.6%] (Figure 2a), and peaking at age 75-84 years old with a striking 40.2% [39.0% to 41.4%] admitted after COVID-19 infection (Figure 2b).

**Figure 2a.**
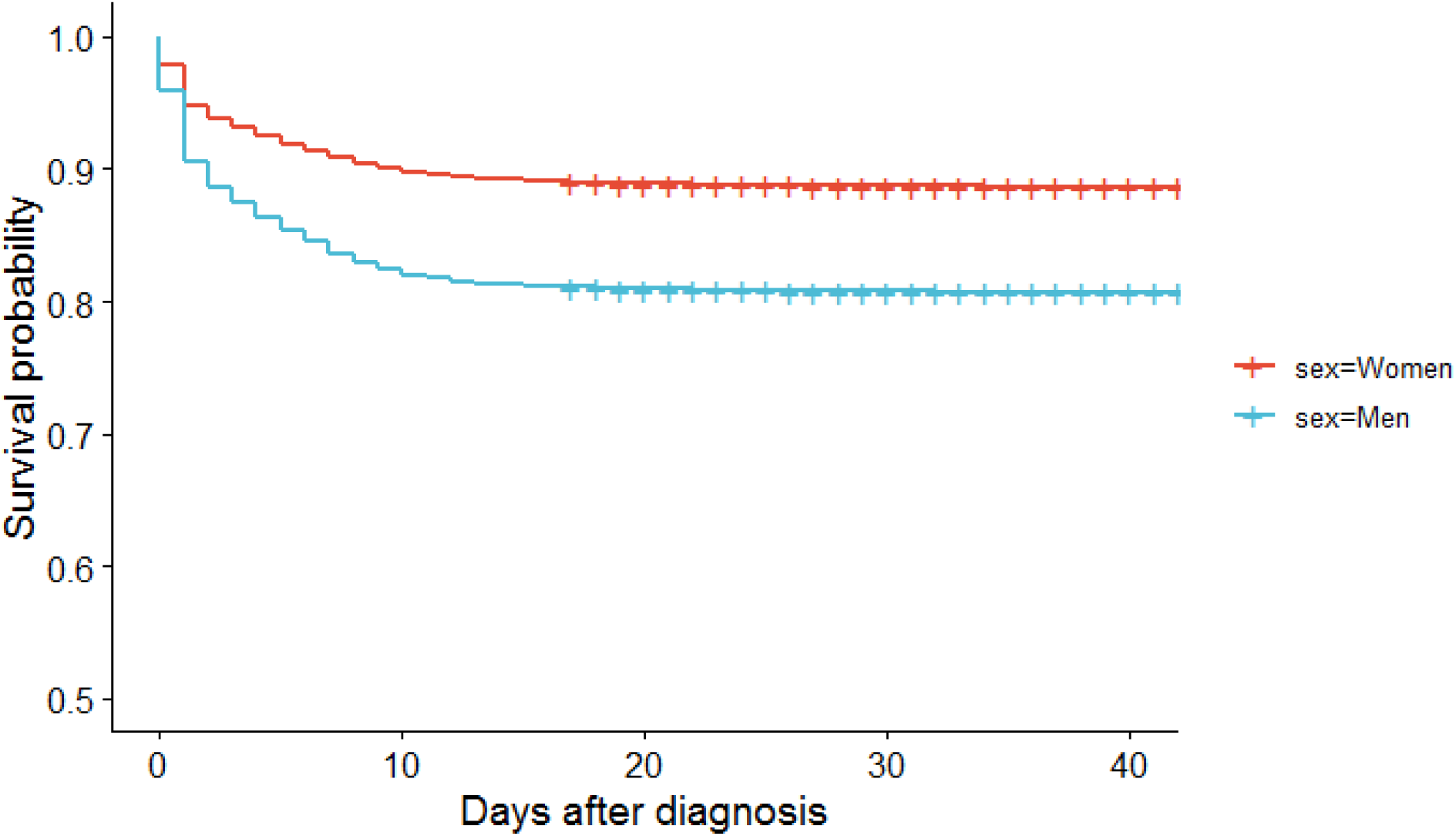
Kaplan-Meier estimates of COVID-19 related hospital admission after COVID-19 diagnosis stratified by sex.

**Figure 2b.**
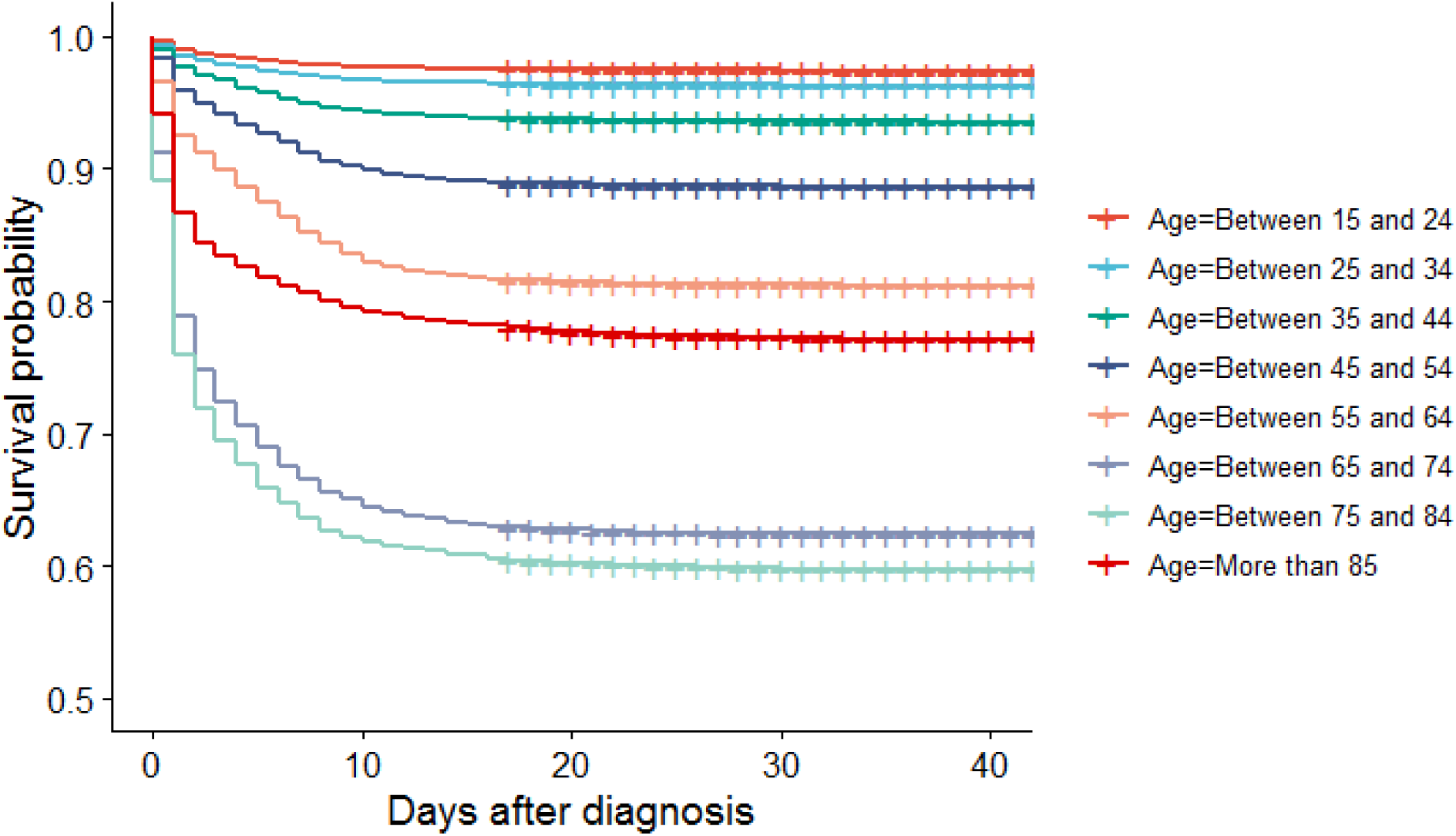
Kaplan-Meier estimates of COVID-19 related hospital admission after COVID-19 diagnosis stratified by age.

The number of deaths observed added to 3,949/98,004, equating to a 30-day fatality of 4.0% [3.9% to 4.2%], higher amongst men (4.8% [4.6% to 5.1%]) than women (3.4% [3.3% to 3.6%]) as seen in Figure 3a. Fatality increased monotonically with age, and more steeply in the elderly: 6.7% [6.2% to 7.3%] in age 65 to 74, 18.2% [17.3% to 19.1%] in age 75 to 84, and up to 29.2% [28.1% to 30.3%] in those aged 85 or older (Figure 3b).

**Figure 3a.**
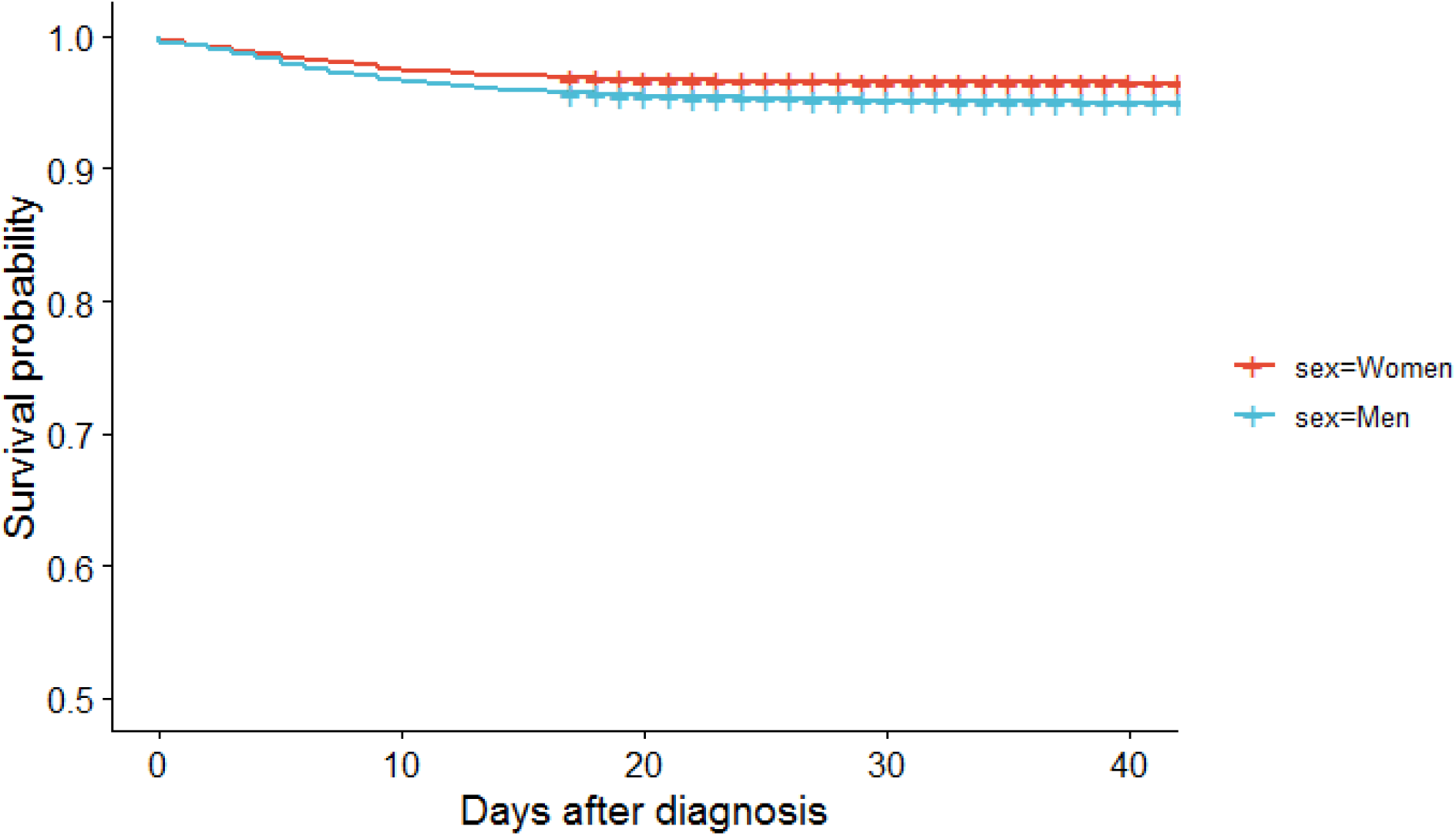
Kaplan-Meier estimates of fatality after COVID-19 diagnosis stratified by sex.

**Figure 3b.**
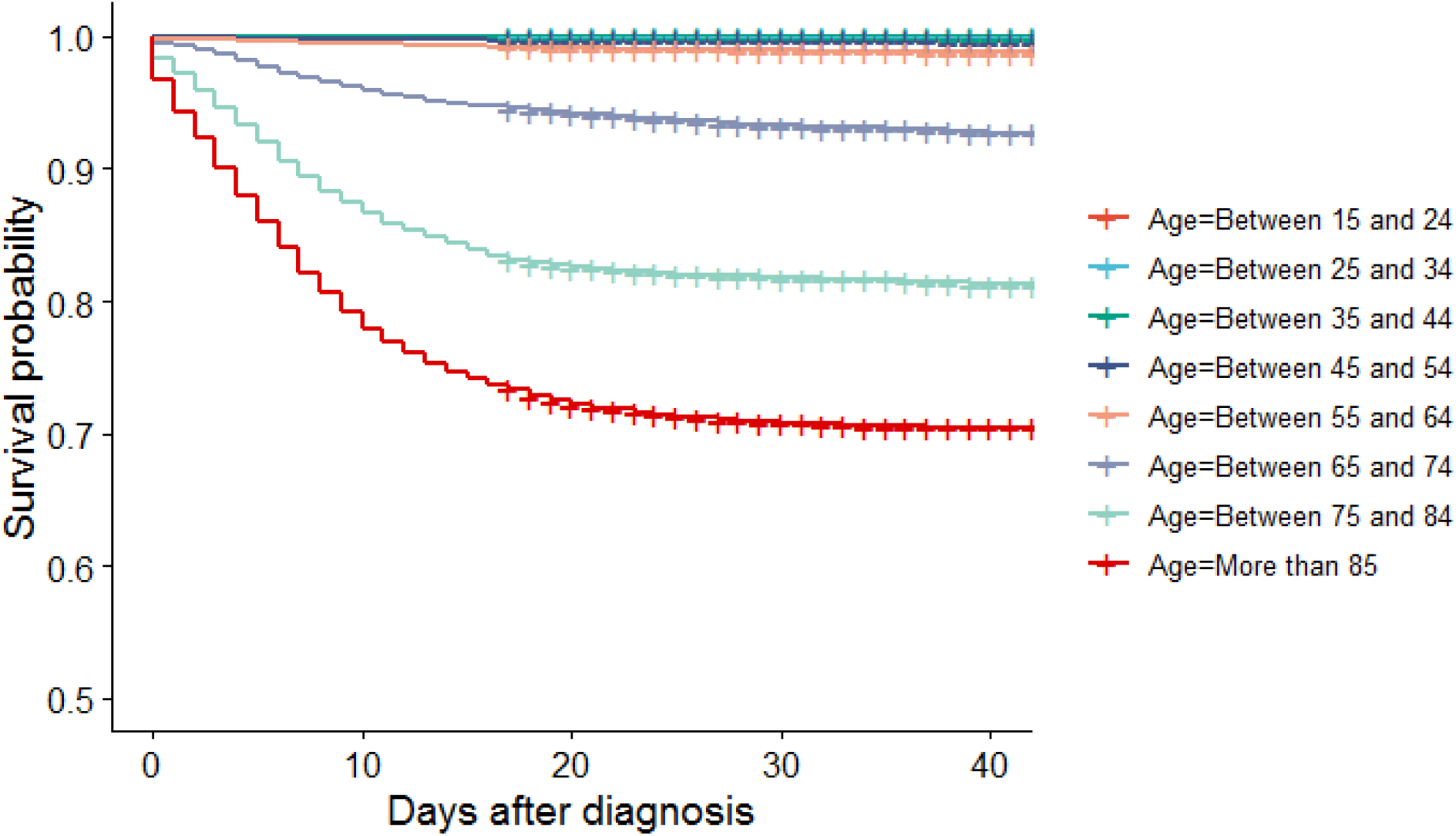
Kaplan-Meier estimates of fatality after COVID-19 diagnosis stratified by age.

Out of the 45,362/46,674 RT-PCR tested patients, those with a positive result had much higher cumulative incidence of hospital admissions (43.1% [42.5% to 43.7%] in RT-PCR+ vs 15.3% [14.5% to 16.1% in RTP-CR-) and a much higher fatality (6.1% [5.8% to 6.4%] in RT-PCR+ vs 0.7% [0.5% to 0.8%] in RT-PCR-).

Finally, outcomes were dramatically different for those residing in nursing homes: although admissions were not much more common than average (16.1% [15.1% to 17.0%]), 30-day fatality was much higher at 25.3% [24.2% to 26.4%].

## Discussion

This is the first study to our knowledge on the characteristics and key health outcomes of COVID-19 amongst patients diagnosed in primary care and outpatient settings. The existence of a universal tax-funded healthcare system and linkage of primary care, hospital, testing and mortality records allowed us to fill a gap in knowledge by providing a comprehensive characterisation of the natural history of COVID-19 including mild as well as severe cases.

We identified a total of 121,263 people diagnosed with COVID-19 between 15th March and 24th April 2020 in Catalonia, most of them initially managed in primary care, with <40% formally tested for SARS-CoV-2 virus. This figure is more than double the official figures, which are based on RT-PCR confirmed cases (48,916 cases confirmed by official sources (https://covid19.isciii.es/) in Catalonia by 30th April 2020. This suggests a much higher proportion of the population have been affected with different levels of severity of the disease, and probably a wider community transmission than previously estimated from official figures.

COVID-19 spread in the whole community, with almost 60% amongst women, and most cases seen in young-middle age adults aged 35-65 years old. Infections were seen in all socio-economic strata in urban environments, and almost 1 in 5 lived in rural areas. Conversely, more severe forms of COVID-19 disease clustered amongst men and elderly people, who account for most hospital admissions and deaths. This is in line with a number of studies in hospitalised COVID-19 populations conducted in the US^12^, China^4^, and Europe^13^, and suggests that age and sex are key risk factors for poor outcomes amongst COVID-19 infectees.

Chronic non-communicable diseases were common amongst COVID-19 infected patients. Probably related to this, most commonly used medicines included analgesics, PPI/antiacids, statins, and anti-hypertensives of different types including ACEi/ARBs used by more than 1 in 10 affected patients. Similar figures have been published elsewhere. Early reports from inpatient cases in China reported a prevalence of diabetes of 7.4%, and of hypertension of 15%^3^. More recently, in an international study including 6,806 hospitalised cases from the US and South Korea^14^, geographical variation was seen in the prevalence of chronic comorbidities, with eg diabetes ranging from a prevalence of 17.9% in South Korea to 43.3% in US veterans and hypertension ranging from 21.8% to 69.7% respectively. Outpatient COVID-19 infectees in our study appear somewhat healthier than US and South Korean inpatient cases, but have a higher prevalence of comorbidities than those reported from China, with diabetes and hypertension prevalences of <10% and <25% respectively. The combination of our data with that obtained from previous studies suggests that the profile of COVID-19 infectees varies in different parts of the world, with no consistent differences seen when outpatient and inpatient cases are compared.

In our data, almost 15% of cases result in a hospital admission in the month after diagnosis in primary care. A preprint of a recent study has compared hospital admissions for influenza compared to inpatient COVID-19, and reported that COVID-19 admitted patients are generally younger and healthier than those admitted for the flu^14^. Additionally, even in this sample including milder forms of disease, 30-day fatality is still high at 4% on average, and almost 5% in men. For comparison, data from 13 previous influenza seasons in Catalonia resulted in an estimated fatality ranging between 0.2% and 0.35%^15^. Previous vaccination^16^ and different age distributions are obvious contributors to this >10-fold lower fatality with influenza compared to COVID-19.

Fatality was much higher (about 1 in 4) in older populations and nursing home residents. In the US, a study in 101 COVID-19 residents of long-term care facilities reported a fatality rate of 33.7%^17^. The spread and severity of COVID-19 shown in this population reflect the recognised vulnerability of nursing home residents to respiratory infections and air-born epidemics including previous coronavirus^18^ and influenza outbreaks and pneumonia^19,20^.

Testing has been limited in primary care settings, even amongst symptomatic patients: only <40% of the identified cases had a RT-PCR taken, representing probably the more severe cases. Positive testing was a clear marker of prognosis, with 43% subsequently hospitalised, and in excess of a 6% 30-day fatality, compared to 15% and <1% respectively amongst clinically diagnosed cases who tested negative for COVID-19. There are currently huge disparities across countries in the implementation of testing following WHO recommendations^21^. Our study demonstrates the value of testing to plan healthcare resource allocation, in addition to its uses to inform public health decision making. Further testing will probably be needed for differential diagnosis with the emergence of effective anti-COVID-19 therapies in the coming _months_^22,23^.

Our study has a number of limitations. First, the use of routinely collected data allowed for the inclusion of a large number of participants, but misclassification is possible as other flu-like or respiratory conditions could potentially be diagnosed as COVID-19 in the context of the current pandemic. However, SIDIAP is a well validated data source^9^ with many previous studies conducted^24^. In a subsample with RT-PCR data available, almost 3 in 4 cases were positive, suggesting that the positive predictive value of primary care diagnosis in our context approached 75%. Given that the other 1 in 4 had better health outcomes (less hospitalizations and lower fatality), it is likely that this misclassification has led to an underestimation of the risk of complications, admissions, and fatality related to COVID-19 infection in our study. Secondly, although we include milder forms of COVID-19 than previous characterisation studies, it is still likely that asymptomatic and cases with little symptoms were advised to stay home and self-isolate to avoid contact and spread in healthcare facilities including primary care practices and hospitals.

This study also has many strengths. The use of primary care records linked to hospital, mortality and testing registers obtained from a universal tax-funded healthcare system allowed for a complete characterisation of the natural history of COVID-19 infection from symptom onset. In addition, the inclusion of COVID-19 cases treated exclusively in outpatient settings (first to date in the literature) allowed for the study of specific subpopulations that are less likely to be admitted to hospital. A striking almost 11,000 COVID-19 cases were identified amongst nursing home residents, more than 9,000 cases had a history of cancer, and almost 600 women were diagnosed with COVID-19 during pregnancy. To date, this is the dataset including the biggest number of COVID-19 cases among these population subgroups known to be more susceptible to infections. For context, previous studies have been limited to <30 cases of COVID-19 amongst patients with prevalent cancer^25,26^. This dataset will therefore be an invaluable resource for studying the effects of SARS-CoV-2 in these populations. Finally, linkage to test data and to comprehensive region-wide hospital and mortality registers allowed us to track COVID-19 infections through a universal healthcare system, whilst avoiding recall bias and loss to follow-up typical of cohort studies.

This is the first study to date on the characteristics, healthcare resource use, and fatality associated with COVID-19 disease as diagnosed in outpatient settings. COVID-19 is often diagnosed and initially managed in primary care, with limited testing leading to under-reporting of cases in official figures, and to an overestimation of fatality. Notwithstanding this, COVID-19 as seen in our study led to hospitalization in 15%, and reached a 30-day fatality of 4%. Our data suggest that although COVID-19 infection has spread in the whole community and most often in young and middle-aged women, severe forms of disease and mortality cluster in older men and amongst nursing home residents. This information is of key relevance for healthcare professionals, public health authorities, and commissioners, now and in future COVID-19 outbreaks.

## Data Availability

No patient level data is directly available from the authors. All the utilised data can be obtained from www.sidiap.org under certain terms and conditions to be consulted with the data custodians.

## Contributors

All authors contributed to the design of the study, the interpretation of the results, and reviewed the manuscript. EC and NM had access to the data, performed the statistical analysis, and acted as guarantors. DP-A, EC, NM, AP-U and TD-S wrote the first draft of the manuscript. All authors critically revised the manuscript. The corresponding author attests that all listed authors meet authorship criteria and that no others meeting the criteria have been omitted.

## Transparency declaration

Lead authors affirm that the manuscript is an honest, accurate, and transparent account of the study being reported; that no important aspects of the study have been omitted; and that any discrepancies from the study as planned have been explained.

## Ethical approval

This study was approved by the Clinical Research Ethics Committee of the IDIAPJGol (project code: 20/070-PCV).

## Acknowledgements

We would like to acknowledge all the patients who suffered from or died of this devastating disease, and their families and careers. We would also like to thank all the healthcare professionals involved in the management of COVID-19 during these challenging times, from primary care to intensive care units in the Catalan healthcare system.

Voldríem reconéixer i tindre un record per tots els pacients que han patit i els què han mort per la COVID-19. Volem també agrair tots els professionals sanitaris que han diagnosticat i tractat aquesta malaltia al sistema català de salut, des dels centres d’atenció primària fins les unitats de cures intensives.

Queremos reconocer y recordar a todos los pacientes que han sufrido y muerto por la COVID-19. Queremos también agradecer a todos los profesionales sanitarios que han diagnosticado y tratado esta enfermedad en el sistema catalán de salud y en España, desde los centros de salud hasta las unidades de cuidados intensivos.

